# Mortality during inpatient admissions for Takotsubo cardiomyopathy: can mortality be predicted with a scoring system?

**DOI:** 10.1101/2022.04.30.22274518

**Authors:** Rohit S. Loomba

## Abstract

**Introduction:** This study aims to identify which comorbidities and demographic characteristics increase the inpatient mortality rate of patients with TCM and to establish a simple scoring system to help identify those at high risk for mortality.

**Methods:** A retrospective cross-sectional study using the Nationwide Inpatient Sample was conducted. Admissions with and without inpatient mortality were then compared. Univariate analyses were conducted to compare admission characteristics with and without inpatient mortality. A logistic regression analysis was conducted with inpatient mortality as the dependent variability and various admission characteristics as the independent variables in order to develop a model to predict inpatient mortality. The model was then used to calculate a score, the TCM mortality score, for each admission. A receiver operator curve analysis was conducted to determine the sensitivity, specificity, negative predictive value, and positive predictive value of the TCM mortality score in prediction of inpatient mortality.

**Results:** A total of 19,454 admissions were included in the final analyses. Inpatient mortality was greater in patients that were older, males, Hispanics or Asian Pacific Islanders, and those with Medicare/Medicaid. In regards to comorbidities, inpatient mortality was greater in those with heart failure, arrhythmia, and acute kidney injury. Those with mortality were also more likely to have had an intra-aortic balloon pump or having had to be placed on extracorporeal membrane oxygenation. A mortality score was found to be 4.4 in the mortality group as compared to 1.6 for those in the non-*mortality* group.

**Conclusion:** Inpatient mortality with Takotsubo cardiomyopathy is approximately 4.0% and those presenting with certain demographic characteristics and comorbidities are at an increased risk for mortality. A simple scoring system can help identify those at high risk for mortality.

## Introduction

Takotsubo cardiomyopathy (TCM) is a syndrome characterized by transient regional systolic dysfunction of the left ventricle. TCM mimics myocardial infarction in some respects; however, angiographic evidence of obstructive coronary artery disease or acute plaque rupture is absent [1]. TCM was first described in Japan in 1990 and has since become a diagnosis reported worldwide. Due to relatively low prevalence the epidemiology of TCM is not entirely elucidated but what has been repeatedly demonstrated is that it is more common in females and the elderly [1,4,13] Otherwise there is a paucity of data regarding factors associated with TCM although it has been hypothesized that genetics, substance abuse, and psychiatric or neurological disorders may increase the risk of TCM.

Mortality in those with TCM appears to be low with most studies reporting a small percentage of patients experiencing inpatient mortality. What mediates the mortality in TCM, however, has not been delineated.

This study aimed to identify patient characteristics and comorbidities that are associated with increased inpatient mortality rate for patients with TCM and to determine if a simple scoring system could be developed to help identify those at high risk for mortality.

## Methods

### Nationwide Inpatient Sample

We conducted a retrospective cross-sectional study using the Nationwide Inpatient Sample. The Nationwide Inpatient Sample, Healthcare Cost Utilization Project (HCUP) by the Agency for Healthcare Research and Quality (AHRQ) is a large database designed to capture data from approximately 20% of all community hospital admissions in the United States. All patients cared for at these selected hospitals are included in the Nationwide Inpatient Sample. Rehabilitation and long-term acute care hospitals are excluded from this database. Patients from all regions of the United States with a variety of payer types are captured in this database. Data from a total of 44 states is captured. A majority of admissions in this database involve adult patients. The Nationwide Inpatient Sample data uses a self-weighting design to help reduce confidence intervals. Data variance from the Nationwide Inpatient Sample can be self-validated for specific iterations.

Institutional reviewer board was not necessary as this study utilized data from a deidentified national database.

### Patient identification

Only admissions with TCM were included in this study and were identified using International Classification of Disease, Ninth edition (ICD-9) code 429.83. A number of comorbidities were then identified in these admissions using the ICD-9 codes that coincided with these diagnoses. Admissions were then segregated into two groups as those who did and did not experience inpatient mortality.

### Statistical analyses

Admissions with and without inpatient mortality were then compared. Univariate analyses were conducted to compare admission characteristics with and without inpatient mortality. Descriptive variables were compared using a chi-square or fisher exact test depending on normalcy of the data while continuous variables were compared using a student t-test or Mann-Whitney-U test depending on the normalcy of the data. Next, a logistic regression analysis was conducted with inpatient mortality as the dependent variability and various admission characteristics as the independent variables in order to develop a model to predict inpatient mortality. Characteristics that were found to be independently associated with increased mortality were included in the model with the coefficient used being the beta-coefficient resulted by the logistic regression.

The model developed to predict inpatient mortality was then used to calculate a score, the TCM mortality score, for each admission. A receiver operator curve analysis was then conducted to determine the sensitivity, specificity, negative predictive value, and positive predictive value of the TCM mortality score in prediction of inpatient mortality.

All statistical analyses were done using SPSS Version 23.0 (IBM corporation, Chicago, IL). Continuous variables are presented as either mean and standard deviation or median and range. Descriptive variables are presented as frequency and percentage. A p-value of less than 0.05 was considered statistically significant.

## Results

### Admission characteristics-univariate analyses

A total of 19,454 admissions were included in the final analyses. Of these 782 (4.0%) experienced inpatient mortality. Inpatient mortality was greater in males (odds ratio 2.5, 95% confidence 2 to 3.3, p< 0.01), Hispanics or Asian Pacific Islanders (p< 0.01), and those with Medicare/Medicaid (p< 0.01). In regards to comorbidities, inpatient mortality was greater in those with heart failure (OR 1.2, 95%CI 1.0 to 1.4, p< 0.01), arrhythmia (OR 1.5, 95%CI 1.2 to 1.8, p< 0.01), and acute kidney injury (OR 4.6, 95%CI 4.0 to 5.4, p< 0.01) (Table 1).

Those with mortality were also more likely to have had an intra-aortic balloon pump (OR 3.7, 95%CI 2.8 to 4.8, p< 0.01) or having had to be placed on extracorporeal membrane oxygenation (OR 88.9, 95%CI 9.9 to 796.0 p< 0.01) (Table 2).

Characteristics independently associated with inpatient mortality-regression analysis Those with inpatient mortality were more likely to be older, with each year of age increasing the odds of inpatient mortality by 1%. Additionally, male gender (OR 1.7, 95%CI 1.5 to 2.5, p< 0.01), Asian/Pacific Islander race (OR 1.9, 95%CI 1.2 to 2.9, p< 0.01), acute kidney injury (OR 3.8, 95%CI 3.2 to 4.5, p< 0.01), arrhythmia (OR 1.2, 95%CI 1.0 to 1.5, p< 0.01), and having Medicaid (OR 1.6, 95%CI 1.1 to 2.2, p< 0.01) were also independently associated with increase in the odds of inpatient mortality (Table 3).

### Takotsubo mortality score

Using the beta-coefficients from the regression analysis a mortality score was obtained by using the following equation: (3 x Male) + (3 x Asian/Pacific Islander) + (6 x Acute kidney injury) + (2 x Medicaid) + (1 x Arrhythmia). The mean TCM mortality score for those in the mortality group was 4.4 compared to 1.6 for those in the non-mortality group (p< 0.01).

Receiver operator curve analysis using the TCM mortality score to predict inpatient mortality score resulted in a curve with an area under the curve of 0.66. The sensitivity, specificity, negative predictive value, and positive predictive value of various TCM mortality scores are demonstrated in table 4. A TCM mortality score of 0 to 5 was associated with a 4.7% risk of inpatient mortality, a score of 5 to 10 was associated with a 6.9% risk of inpatient mortality, and a score of greater than 10 was associated with a 22.7% risk of inpatient mortality (Table 5).

## Discussion

To our knowledge, this is the largest study of TCM mortality until date. Our study demonstrates an inpatient mortality rate of 4.0% in those with TCM. Independent factors associated with inpatient mortality included male gender, Asian/Pacific Islander race, acute kidney injury, arrhythmia, and having Medicaid.

Previous studies have demonstrated that TCM is more frequently noted in females. 89.9 percent of patients with TCM were found to be female in the International Takotsubo Registry. The mean age was 66.4 years [13]. Previous studies have also noted this trend and attributed it to an increased prevalence of cardiogenic shock, cardiac arrest, and respiratory failure [7]. Female TCM patients have been shown to have higher rates of emotional stress to triggers such as unexpected death of relatives, catastrophic medical diagnoses, or domestic argument/abuse when compared to their male counterparts which may somehow mediate the risk of mortality. When compared to females, males are more likely to develop TCM as a result to physical stressors such as acute respiratory failure, central nervous system condition, or infection [14].

The event triggering TCM, whether emotional versus physical, may alter pathophysiology of the disease. It is possible that the greater the stress response to the inciting event the greater the ventricular dysfunction that results from it. This has yet to be demonstrated. Additionally, most females with TCM are post-menopausal and thus have lost the cardioprotective benefits of estrogen [8a]. Estrogen is a strong cardioprotective hormone and postmenopausal females have lower estrogen levels than males of a similar age. Females may be more likely to present with TCM because males with TCM tend to die at earlier stages of disease progression sometimes before TCM has been detected thus leading to a failure of diagnosis [11, 12, 14].

Our study also demonstrated that mortality was different in those with different races. Asian/Pacific islanders had the greatest risk of mortality with 7.8% mortality. Current medical literature acknowledges electrocardiographic differences between Caucasians suffering from TCM and asians. Caucasians are more likely to have T-wave inversion on ECG, whereas Asians more often have ST-segment elevation [5,13]. A recent study has also found ST elevation to have a major association with major cardiac events at follow-up [10]. Additionally, T-wave inversion was more common than ST-elevation in African American patients [5]. The difference in T-wave findings may imply that there is a degree of ischemia that occurs in the setting of TCM and that somehow this differs by race, although the underlying mechanism of this cannot be inferred with the current data.

Acute kidney injury and arrhythmia were the two comorbidities identified to be associated with inpatient mortality. Acute kidney injury has been demonstrated in several studies to be associated with increased risk in a number of different inpatient populations. Arrhythmia was also associated with increased mortality and has been demonstrated to have the same impact in a variety of other inpatient populations as well.

Those with Medicaid were also demonstrated to have a greater risk of inpatient mortality. This has been demonstrated in previous patient populations with several hypotheses being proposed as to why such differences may exist [2,3,6].

We also aimed to model inpatient mortality to develop a TCM mortality score that could be used to identify risk of mortality in inpatients with TCM. While the model had an area under the curve of 0.66, scores did differ between the mortality and non-mortality groups and increasing scores were associated with increased risk of mortality. Earlier in this manuscript we described the predicted mortality with three different score strata. The strata with the lowest scores had an inpatient mortality of 4.7%, similar to the overall rate of 4.0%. The middle strata had an inpatient mortality of 6.9% and the highest strata had an inpatient mortality of 22.7%. We feel that this scoring system can help identify higher risk patients despite the low area under the curve of the overall model until a more accurate model is developed.

While our study offers several novel insights into characteristics of TCM there are limitations. Limitations of this study stem from the nature of the study and its utilization of NIS data. Firstly, this was a retrospective study based on data regarding admissions. Admissions could not be identified as readmissions or being for the same patient, thus some patients are undoubtedly represented by multiple admissions in the analyses but the impact of this cannot be quantified. Also, the NIS database utilizes data captured by ICD9 coding. There may certainly be inaccuracies and oversights with coding and thus this could introduce a discrepancy between the actual and represented frequencies of various characteristics and comorbidities. Specifically, frequencies in our study are likely underestimations. An additional limitation of this study is the absence of laboratory data.

## Conclusion

Inpatient mortality with Takotsubo cardiomyopathy is approximately 4.0%. Inpatients with Takotsubo cardiomyopathy are at increased risk for mortality with the presence of certain demographic characteristics and comorbidities. A simple scoring system can help identify those at high risk for mortality.

## Data Availability

All data produced in the present work are contained in the manuscript

## References

[1] Akashi, Yoshihiro J., David S. Goldstein, Giuseppe Barbaro, and Takashi Ueyama. “Takotsubo Cardiomyopathy: A New Form of Acute, Reversible Heart Failure.” Circulation 118, no. 25 (December 16, 2008): 2754–62. https://doi.org/10.1161/CIRCULATIONAHA.108.767012.

[2] Ayanian, John Z., Betsy A. Kohler, Toshi Abe, and Arnold M. Epstein. “The Relation between Health Insurance Coverage and Clinical Outcomes among Women with Breast Cancer.” New England Journal of Medicine 329, no. 5 (July 29, 1993): 326–31. https://doi.org/10.1056/NEJM199307293290507.

[3] Burstin, Helen R., Stuart R. Lipsitz, and Troyen A. Brennan. “Socioeconomic Status and Risk for Substandard Medical Care.” JAMA 268, no. 17 (November 4, 1992): 2383–87. https://doi.org/10.1001/jama.1992.03490170055025.

[4] Donohue, Daniel, and Mohammad-Reza Movahed. “Clinical Characteristics, Demographics and Prognosis of Transient Left Ventricular Apical Ballooning Syndrome.” Heart Failure Reviews 10, no. 4 (December 1, 2005): 311–16. https://doi.org/10.1007/s10741-005-8555-8.

[5] Franco, Emiliana, Andre Dias, Nikoloz Koshkelashvili, Gregg S. Pressman, Kathy Hebert, and Vincent M. Figueredo. “Distinctive Electrocardiographic Features in African Americans Diagnosed with Takotsubo Cardiomyopathy.” Annals of Noninvasive Electrocardiology 21, no. 5 (September 1, 2016): 486–92. https://doi.org/10.1111/anec.12337.

[6] Hasan, Omar, E. John Orav, and LeRoi S. Hicks. “Insurance Status and Hospital Care for Myocardial Infarction, Stroke, and Pneumonia.” Journal of Hospital Medicine 5, no. 8 (October 2010): 452–59. https://doi.org/10.1002/jhm.687.

[7] Khera R, Light-McGroary K, Zahr F, Horwitz PA, Girotra S. Trends in Hospitalization for Takotsubo Cardiomyopathy in the United States. American heart journal. 2016;172:53–63. doi:10.1016/j.ahj.2015.10.022.

[8] Madhavan, M., and A. Prasad. “Proposed Mayo Clinic Criteria for the Diagnosis of Tako-Tsubo Cardiomyopathy and Long-Term Prognosis.” Herz 35, no. 4 (June 1, 2010): 240–44. https://doi.org/10.1007/s00059-010-3339-x.

[9] Murakami, Tsutomu, Tsutomu Yoshikawa, Yuichiro Maekawa, Tetsuro Ueda, Toshiaki Isogai, Konomi Sakata, Ken Nagao, Takeshi Yamamoto, and Morimasa Takayama. “Gender Differences in Patients with Takotsubo Cardiomyopathy: Multi-Center Registry from Tokyo CCU Network.” PLoS ONE 10, no. 8 (August 28, 2015). https://doi.org/10.1371/journal.pone.0136655.

[10] Santoro, Francesco, Thomas Stiermaier, Nicola Tarantino, Francesca Guastafierro, Tobias Graf, Christian Möller, Luigi F. M. Di Martino, et al. “Impact of Persistent ST Elevation on Outcome in Patients with Takotsubo Syndrome. Results from the GErman Italian STress Cardiomyopathy (GEIST) Registry.” International Journal of Cardiology, November 22, 2017. https://doi.org/10.1016/j.ijcard.2017.11.068.

[11] Schneider, Birke, Anastasios Athanasiadis, Claudia Stöllberger, Wolfgang Pistner, Johannes Schwab, Uta Gottwald, Ralph Schoeller, et al. “Gender Differences in the Manifestation of Tako-Tsubo Cardiomyopathy.” International Journal of Cardiology 166, no. 3 (July 1, 2013): 584–88. https://doi.org/10.1016/j.ijcard.2011.11.027.

[12] Stöllberger, Claudia, and Josef Finsterer. “Why Does Takotsubo (‘Broken Heart Syndrome’) Affect More Females than Males?” International Journal of Cardiology 147, no. 1 (February 17, 2011): 175–76. https://doi.org/10.1016/j.ijcard.2010.12.026.

[13] Templin, Christian, Jelena R. Ghadri, Johanna Diekmann, L. Christian Napp, Dana R. Bataiosu, Milosz Jaguszewski, Victoria L. Cammann, et al. “Clinical Features and Outcomes of Takotsubo (Stress) Cardiomyopathy.” New England Journal of Medicine 373, no. 10 (September 3, 2015): 929–38. https://doi.org/10.1056/NEJMoa1406761.

[14] Weidner, K. J., I. El-Battrawy, M. Behnes, K. Schramm, C. Fastner, J. Kuschyk, U. Hoffmann, U. Ansari, M. Borggrefe, and I. Akin. “Sex Differences of In-Hospital Outcome and Long-Term Mortality in Patients with Takotsubo Cardiomyopathy.” Therapeutics and Clinical Risk Management, July 12, 2017. https://doi.org/10.2147/TCRM.S131760.

