## Supplementary material for "Mortality during inpatient admissions for Takotsubo cardiomyopathy: can mortality be predicted with a scoring system?": Table 1

|  | **No mortality**  **(n= 18,672)** | **Mortality**  **(n= 782)** | **Odds ratio (95% confidence interval)** | **p-value** |
| --- | --- | --- | --- | --- |
| ***Female*** | 16,541 (88.6) | 613 (78.4) | 0.4 (0.3 to 0.5) | < 0.01 |
| ***Race***  ***Caucasian***  ***African American***  ***Hispanic***  ***Asian/pacific islander***  ***Native American***  ***Other*** | 13,358 (83.0)  1,080 (6.7)  857 (5.3)  294 (1.8)  101 (0.6)  407 (2.5) | 536 (78.0)  50 (7.3)  46 (6.7)  25 (3.6)  *** (***)  27 (3.9) | -- | < 0.01 |
| ***Expected payer***  ***Medicare***  ***Medicaid***  ***Private insurance***  ***Self-pay***  ***No charge***  ***Other*** | 11,327 (60.7)  1,237 (6.6)  4,885 (26.2)  716 (3.8)  54 (0.3)  429 (2.3) | 511 (65.4)  76 (9.7)  149 (19.1)  31 (4.0)  *** (***)  13 (1.7) | -- | < 0.01 |
| ***Weekend admission*** | 4,334 (23.2) | 191 (24.2) | 1.0 (0.9 to 1.2) | 0.43 |
| ***Admission month***  ***January***  ***February***  ***March***  ***April***  ***May***  ***June***  ***July***  ***August***  ***September***  ***October***  ***November***  ***December*** | 1,196 (6.8)  1,192 (6.8)  1,373 (7.8)  1,329 (7.6)  1,445 (8.2)  1,388 (7.9)  1,501 (8.5)  1,642 (9.3)  1,646 (9.4)  1,710 (9.7)  1,610 (9.2)  1,540 (8.8) | 57 (7.6)  61 (8.1)  50 (6.7)  51 (6.8)  63 (8.4)  62 (8.3)  74 (9.9)  62 (8.3)  60 (8.0)  63 (8.4)  77 (10.3)  70 (9.3) | -- | 0.45 |
| ***Obesity*** | 1,307 (7.0) | 19 (2.4) | 0.3 (0.2 to 0.5) | < 0.01 |
| ***Heart failure*** | 6,316 (33.8) | 300 (38.4) | 1.2 (1.0 to 1.4) | < 0.01 |
| ***Hypertension*** | 9,665 (51.8) | 246 (31.5) | 0.4 (0.3 to 0.5) | < 0.01 |
| ***Arrhythmia*** | 2,990 (16.0) | 176 (22.5) | 1.5 (1.2 to 1.8) | < 0.01 |
| ***Diabetes mellitus*** | 3,519 (18.8) | 99 (12.7) | 0.6 (0.5 to 0.7) | < 0.01 |
| ***Acute kidney injury*** | 2,152 (11.5) | 296 (37.9) | 4.6 (4.0 to 5.4) | < 0.01 |
| ***Chronic kidney disease*** | 1,515 (8.1) | 70 (9.0) | 1.1 (0.8 to 1.4) | 0.40 |
| ***Current smoking*** | 2,581 (13.8) | 60 (7.7) | 0.5 (0.3 to 0.7) | < 0.01 |
| ***Takotsubo mortality score*** | 1.6 | 4.4 | -- | < 0.01 |
