## Supplementary material for "Mortality during inpatient admissions for Takotsubo cardiomyopathy: can mortality be predicted with a scoring system?": table 2

|  | **No mortality**  **(n= 18,672)** | **Mortality**  **(n= 782)** | **Odds ratio (95% confidence interval)** | **p-value** |
| --- | --- | --- | --- | --- |
| ***AICD insertion*** | 119 (0.7) | *** (***) | *** | 0.02 |
| ***IABP*** | 395 (2.5) | 62 (8.6) | 3.7 (2.8 to 4.8) | < 0.01 |
| ***ECMO*** | *** (***) | *** (***) | 88.9 (9.9 to 796.0) | < 0.01 |
