## Supplementary material for "Mortality during inpatient admissions for Takotsubo cardiomyopathy: can mortality be predicted with a scoring system?": table 3

| **Factors independently associated with Takotsubo cardiomyopathy** | |
| --- | --- |
| - Older age | Each increase in age by 1 year increases mortality by mortality by 1% |
| - Male gender | OR 1.7, 95%CI 1.4 to 2.5, p <0.01 |
| - Asian/Pacific Islander race | OR 1.9, 95%CI 1.2 to 2.9, p< 0.01 |
| - Acute kidney injury | OR 3.8, 95%CI 3.2 to 4.5, p< 0.01 |
| - Arrhythmia | OR 1.2, 95%CI 1.0 to 1.5, p< 0.01 |
| - Medicaid | OR 1.6, 95%CI 1.1 to 2.2, p< 0.01 |
