## Supplementary material for "Mortality during inpatient admissions for Takotsubo cardiomyopathy: can mortality be predicted with a scoring system?": table 4

| **Mortality score** | **Sensitivity** | **Specificity** | **Negative predictive value** | **Positive predictive value** |
| --- | --- | --- | --- | --- |
| ***1.0*** | 64% | 63% | 72% | 53% |
| ***5.0*** | 39% | 84% | 67% | 61% |
| ***9.0*** | 16% | 94% | 62% | 64% |
| ***11.0*** | 7% | 97% | 61% | 61% |
| ***13.0*** | 4% | 98% | 60% | 57% |
